# Does household income predict health and educational outcomes in childhood better than neighbourhood deprivation?

**DOI:** 10.1101/2024.07.25.24310986

**Authors:** Ieva Skarda, Richard Cookson, Ruth Gilbert

**Author notes:** Address correspondence to Ieva Skarda. All authors contributed to the conceptualisation, methods and writing of iterative drafts. Ieva Skarda conducted the analyses, with input from Richard Cookson and Ruth Gilbert.

## Abstract

**Background:** Public health research and prevention policies often use the small area Index of Multiple Deprivation (IMD) at neighbourhood level to proxy individual socio-economic status because it is readily available. We investigated what household income adds to IMD in early childhood for predicting adverse health in adolescence.

**Methods:** Using data from the Millennium Cohort Study, we analysed IMD and self-reported equivalised household income (ages 0-5) to predict outcomes at age 17: poor academic achievement, psychological distress, poor health, smoking, and obesity. Predictions were compared using IMD quintiles alone, household income alone, and both together.

**Results:** Household income was a stronger and more consistent predictor of age 17 outcomes than IMD and revealed inequalities within neighbourhoods. Decreasing household income showed steep gradients in educational attainment and smoking across all IMD quintiles, and moderate gradients in obesity, psychological distress, and poor health in most quintiles. IMD did not predict smoking or psychological distress within any income group, or educational attainment within the poorest income group.

**Conclusion:** Household income is associated with inequality gradients within all quintiles of neighbourhood IMD. Early childhood public health strategies should consider household income in combination with neighbourhood deprivation.

## Introduction

The Index of Multiple Deprivation (IMD) has been widely used as a neighbourhood measure of deprivation over the last two decades to guide UK policy on health disparities (see Table A1 in the appendix for details). The success of IMD is due, in part, to its ready availability and routine linkage to administrative health and other datasets. Furthermore, IMD consistently shows a clear and monotonic social gradient in almost all indicators of health which underpins its use as a tool for identifying need and allocating services locally and nationally.^[1]^

Household income is an important indicator of socio-economic status that is strongly associated with health^[2–9]^ yet is challenging to measure and link to routine health data.^[10]^ Hence, public health research and prevention policies often use the IMD at the neighbourhood level to proxy individual socio-economic status. For example, NHS England statistics aimed at reducing health inequalities in children and young people currently focus on the most deprived 20% of neighbourhoods (in addition to vulnerable groups identified locally, such as ethnic minorities, those with learning disability, care leavers and those with five specific health conditions).^[11]^

However, IMD is a blunt instrument for guiding policies to reduce health inequalities. Firstly, 55% to 62% of the poorest households, based on household income, live outside the most deprived 20% of neighbourhoods (ranked using IMD), so are likely to be missed by such policies.^[12–14]^ Secondly, focusing on the most deprived ignores the gradient of adverse health outcomes across the whole distribution of deprivation, particularly income deprivation. Thirdly, IMD does not capture the fact that household income is highly amenable to policy interventions directly affecting income, such as on wages, benefits, wealth, housing, and costs of other essentials.

This raises the question of whether household income, grouped in a useful way that avoids risk of disclosure, could add value to existing neighbourhood measures of deprivation.

Few studies have directly assessed the added value of individual-level measures of income for predicting adverse health outcomes – such as household income or personal earnings – compared with neighbourhood-level measures of deprivation. A recent scoping review concluded that individual-level measures of social disadvantage tended to identify stronger associations with adverse child health than neighbourhood-level measures.^[15]^ More research is needed on the advantages of combining individual-level and neighbourhood-level measures, including to address intersectional inequalities and to counter the problem of the ecological fallacy inherent in neighbourhood-level measures.

Our aim was to address the evidence gap on whether household income adds important information, separately and in combination with neighbourhood IMD, that could be used by services and policy makers to identify groups in childhood with poor long-term health outcomes who might benefit from early intervention. We used the Millennium Cohort Study (MCS), which contains parent-reported household income and IMD in the early childhood years. We explored associations between quintiles of neighbourhood IMD and household income alone, and in combination, with five health-related outcomes at age 17: poor educational attainment, psychological distress, poor health, smoking, and obesity. All five outcomes have strong evidence of long-term impacts on health in adulthood.^[16]^ We compared associations between these outcomes and household income and neighbourhood deprivation separately and assessed the added value of both variables combined.

## Methods

### Data and sample

Data for this study were derived from waves 1 to 3 and wave 7 of the MCS, a nationally representative retrospective cohort study following individuals born in the UK between September 1, 2000, and January 11, 2002. The first survey in 2001–02 included 18,819 children, with subsequent surveys at ages 3, 5, 7, 11, 14, and 17. Additional details on MCS can be found elsewhere.^[17]^ Ethics approval was obtained through the National Health Service Research Ethics Committee system, with written informed consent from parents up to age 14 and verbal consent from those aged 16 and older.

We addressed missing data primarily due to attrition using multiple imputation with chained equations. Imputation was carried out from the age 3 survey (sweep 2), our primary imputation sweep, which included 1,389 additional families not surveyed in sweep 1.^[18], [19]^ Our primary analysis dataset includes observations on 15,367 children after imputation. Analyses were weighted using inverse probability weights to adjust for attrition and sampling design. Further details on the derivation of our primary analysis dataset are in the appendix (section A1 and Figure A1)."

### Measures

Data on neighbourhood deprivation was based on the IMD 2004 Overall Deciles (ranked separately for England, Scotland, Wales and Northern Ireland) which are geographically linked to the MCS households at the lower super output level (LSOA; see table A1, appendix, for details on all variables used in the analyses). The decile group numbers at the first three waves (9 months, 3 years, 5 years) were averaged over these waves, rounded and transformed into quintile groups.

Between 2001 and 2006, questions about household income in early childhood (after tax and other deductions but before housing costs) were asked during home interviews and reported by the main parent/ caregiver at ages 9 months, 3 years, and 5 years. Imputed banded responses were converted into continuous values and equivalised based on household size and composition.^[20]^ Average household income across the first three waves was ranked into income quintiles. Health-related outcomes at age 17 are summarized in the appendix (Table A1) and detailed elsewhere.^[16]^

### Statistical analyses

We cross-tabulated the percentage prevalence of each of the five adverse health-related outcomes at age 17 across the five IMD quintiles and the five income quintiles in early childhood, as well as the twenty-five sub-groups defined by both IMD and income quintiles, using heatmap shading to visualize the degree of outcome prevalence (Table 1). We also report how the sample children were distributed across these subgroups (appendix Figure A2 and Figure A3).

**Table 1.** Crosstabulation heatmap of incidence of adverse adolescent outcomes at age 17 in the different IMD and income quintile subgroups.

|  | Most deprived<br>20% IMD | Most deprived<br>20-40% IMD | Middle<br>40-60% IMD | Least deprived<br>60-80% IMD | Least deprived<br>80-100% IMD | Total |
| --- | --- | --- | --- | --- | --- | --- |
| *Poor academic achievement, %* |  |  |  |  |  |  |
| Poorest 20% income | 61.9<br>[59.4-64.5] | 63.6<br>[58.9-68.3] | 64.5<br>[58.2-70.8] | 58.8<br>[47.5-70] | 61.9<br>[45.1-78.7] | 62.6<br>[60.4-64.7] |
| Poorest 20-40% income | 53.1<br>[49.7-56.5] | 48.9<br>[44.7-53.1] | 48.4<br>[43.7-53.1] | 45.1<br>[38.6-51.6] | 39.9<br>[30.7-49.1] | 49.1<br>[47-51.2] |
| Middle 40-60% income | 41.3<br>[36.6-46] | 37.3<br>[32.8-41.8] | 35.8<br>[31.6-40.1] | 31.2<br>[26.4-36.1] | 23.1<br>[17.6-28.5] | 34<br>[32-36.1] |
| Richest 60-80% income | 33.8<br>[26.6-40.9] | 29.9<br>[25.4-34.4] | 28.1<br>[24-32.1] | 21.1<br>[17.4-24.9] | 18.5<br>[14.5-22.4] | 24.3<br>[22.3-26.3] |
| Richest 80-100% income | 11.2<br>[0.8-21.5] | 15.7<br>[10.6-20.7] | 17.6<br>[13.3-21.9] | 14.9<br>[11.9-17.9] | 11.2<br>[9-13.3] | 13.9<br>[12.3-15.4] |
| **Total** | **54.2**<br>**[52.2-56.1]** | **44.3**<br>**[42-46.6]** | **37.6**<br>**[35.5-39.8]** | **26.5**<br>**[24.4-28.6]** | **18.4**<br>**[16.4-20.5]** | **36.8**<br>**[35.8-37.8]** |
| *Psychological distress, %* |  |  |  |  |  |  |
| Poorest 20% income | 16.2<br>[13.8-18.6] | 19.2<br>[15.9-22.4] | 22.1<br>[17.1-27.1] | 19.3<br>[10.6-27.9] | 24.2<br>[8.5-39.9] | 18.2<br>[16.3-20.1] |
| Poorest 20-40% income | 16.2<br>[13.5-19] | 18.9<br>[15.9-21.9] | 17.4<br>[13.9-20.9] | 17.1<br>[11.4-22.8] | 14.2<br>[6.8-21.6] | 17.2<br>[15.6-18.9] |
| Middle 40-60% income | 16<br>[12.5-19.6] | 13.1<br>[10.3-16] | 16<br>[13-19.1] | 17.4<br>[13.8-21] | 16.6<br>[11.9-21.3] | 15.7<br>[14.2-17.3] |
| Richest 60-80% income | 18.9<br>[12.4-25.5] | 13.6<br>[9.9-17.3] | 16<br>[12.6-19.4] | 11.5<br>[8.6-14.4] | 12.6<br>[9.4-15.8] | 13.7<br>[12-15.3] |
| Richest 80-100% income | 11.9<br>[3.9-20] | 13.5<br>[8.9-18.1] | 12.6<br>[9.3-15.8] | 11.3<br>[8.7-13.9] | 11.2<br>[8.9-13.5] | 11.7<br>[10.2-13.1] |
| **Total** | **16.2**<br>**[14.7-17.8]** | **16.3**<br>**[14.8-17.9]** | **16.6**<br>**[15.1-18.1]** | **13.9**<br>**[12.2-15.5]** | **13**<br>**[11.2-14.9]** | **15.3**<br>**[14.6-16]** |
| *Poor health, %* |  |  |  |  |  |  |
| Poorest 20% income | 13.5<br>[11.4-15.7] | 12<br>[8.9-15.1] | 14.8<br>[10.5-19.2] | 8.7<br>[1.9-15.4] | 13.8<br>[2.5-25.2] | 13.1<br>[11.3-14.8] |
| Poorest 20-40% income | 10.4<br>[8-12.7] | 10.8<br>[8.3-13.4] | 7.1<br>[4.2-10] | 8.6<br>[4.4-12.8] | 5.6 [0.5-10.7] | 9.2<br>[7.9-10.6] |
| Middle 40-60% income | 9.8<br>[6.4-13.1] | 8.3<br>[5.7-10.9] | 6.4<br>[4.1-8.6] | 6.4<br>[3.5-9.3] | 5.2<br>[2.3-8.2] | 7.1<br>[5.9-8.3] |
| Richest 60-80% income | 5.6<br>[1.1-10] | 7.7<br>[5-10.3] | 8.6<br>[6.2-11] | 4.8<br>[2.8-6.7] | 4.2<br>[2.2-6.2] | 6.1<br>[5-7.2] |
| Richest 80-100% income | 5.2<br>[-1.1-11.5] | 5.8<br>[2.5-9.2] | 5.5<br>[3.3-7.7] | 3.8<br>[2.2-5.3] | 3.5<br>[2.1-4.9] | 4.2<br>[3.2-5.1] |
| **Total** | **11.5**<br>**[10-13]** | **9.6**<br>**[8.3-11]** | **8.1**<br>**[6.8-9.3]** | **5.5**<br>**[4.2-6.8]** | **4.4**<br>**[3.4-5.4]** | **7.9**<br>**[7.3-8.5]** |

| *Smoking, %* |  |  |  |  |  |  |
| --- | --- | --- | --- | --- | --- | --- |
| Poorest 20% income | 16.7<br>[14.4-18.9] | 18.6<br>[15.2-21.9] | 19.7<br>[14.6-24.9] | 17.5<br>[8.7-26.4] | 26.8<br>[9.5-44.1] | 17.9<br>[16.1-19.7] |
| Poorest 20-40% income | 13.9<br>[11.5-16.3] | 14.2<br>[11-17.3] | 11.3<br>[7.9-14.7] | 12.6<br>[7.9-17.3] | 11.4<br>[5.5-17.3] | 13.1<br>[11.6-14.5] |
| Middle 40-60% income | 11<br>[7.5-14.4] | 8<br>[5.6-10.5] | 10.1<br>[7.4-12.8] | 7.6<br>[4.5-10.6] | 7.7<br>[4.3-11.1] | 8.8<br>[7.5-10.2] |
| Richest 60-80% income | 7.3<br>[2.5-12.1] | 7.1<br>[4.2-9.9] | 7.1<br>[4.8-9.4] | 5.7<br>[3.7-7.7] | 6.4<br>[4.2-8.6] | 6.5<br>[5.4-7.6] |
| Richest 80-100% income | 4.3<br>[-2.9-11.5] | 5.2<br>[1.8-8.5] | 7<br>[4.4-9.6] | 5.1<br>[3.1-7.1] | 4.3<br>[2.8-5.7] | 5.1<br>[4-6.2] |
| **Total** | **14.4**<br>[12.8-15.9] | **12.0**<br>[10.6-13.4] | **10.5**<br>[9.1-11.9] | **7.5**<br>[6.1-8.8] | **6.5**<br>[5.3-7.8] | **10.3**<br>[9.7-10.9] |

| *Obesity, %* |  |  |  |  |  |  |
| --- | --- | --- | --- | --- | --- | --- |
| Poorest 20% income | 25<br>[22.5-27.4] | 22.8<br>[19.4-26.2] | 17.7<br>[12.6-22.7] | 23.3<br>[14.7-31.9] | 13.5<br>[1.6-25.3] | 23.0<br>[21.2-24.7] |
| Poorest 20-40% income | 21.3<br>[18.6-24.1] | 22.5<br>[19.1-25.8] | 20.4<br>[16.3-24.4] | 22.6<br>[16.5-28.7] | 24.6<br>[16.2-33] | 21.8<br>[19.9-23.6] |
| Middle 40-60% income | 24.9<br>[20.6-29.2] | 22.3<br>[18.9-25.7] | 18.6<br>[15.2-22] | 15.2<br>[11.3-19] | 15<br>[10.4-19.5] | 19.1<br>[17.3-20.8] |
| Richest 60-80% income | 24.9<br>[18.7-31.2] | 20.8<br>[17.1-24.6] | 17.4<br>[14.3-20.5] | 15.5<br>[12.6-18.5] | 14.3<br>[11.1-17.5] | 17.1<br>[15.5-18.6] |
| Richest 80-100% income | 14.2<br>[5.0-23.4] | 20.8<br>[15.2-26.3] | 13.7<br>[10.2-17.1] | 14.6<br>[11.8-17.4] | 8.9<br>[6.8-11] | 12.6<br>[11.1-14] |
| **Total** | **23.7**<br>[22-25.4] | **22.1**<br>[20.4-23.9] | **17.8**<br>[16.1-19.4] | **16.5**<br>[14.7-18.3] | **12.6**<br>[11-14.2] | **18.7**<br>[17.9-19.5] |

*Note: 95% confidence intervals reported in the square brackets. The heatmap shading visualises the magnitude of outcome prevalence, such that a darker shade corresponds to a higher prevalence and a lighter shade – to a lower prevalence.*

We plotted the percentage prevalence of adverse outcomes across the sub-groups in two ways: (1) sorting first by IMD quintile and then by income quintile, allowing us to observe the inequality gradient within each IMD quintile as income decreases (Figure 1), and (2) sorting first by income quintile and then by IMD quintile (Figure 2), allowing us to observe the inequality gradient within each income quintile as IMD decreases. The aim was to determine whether the inequality gradient changes when examining within-group variation and if this varies depending on the measure used.

**Figure 1:**
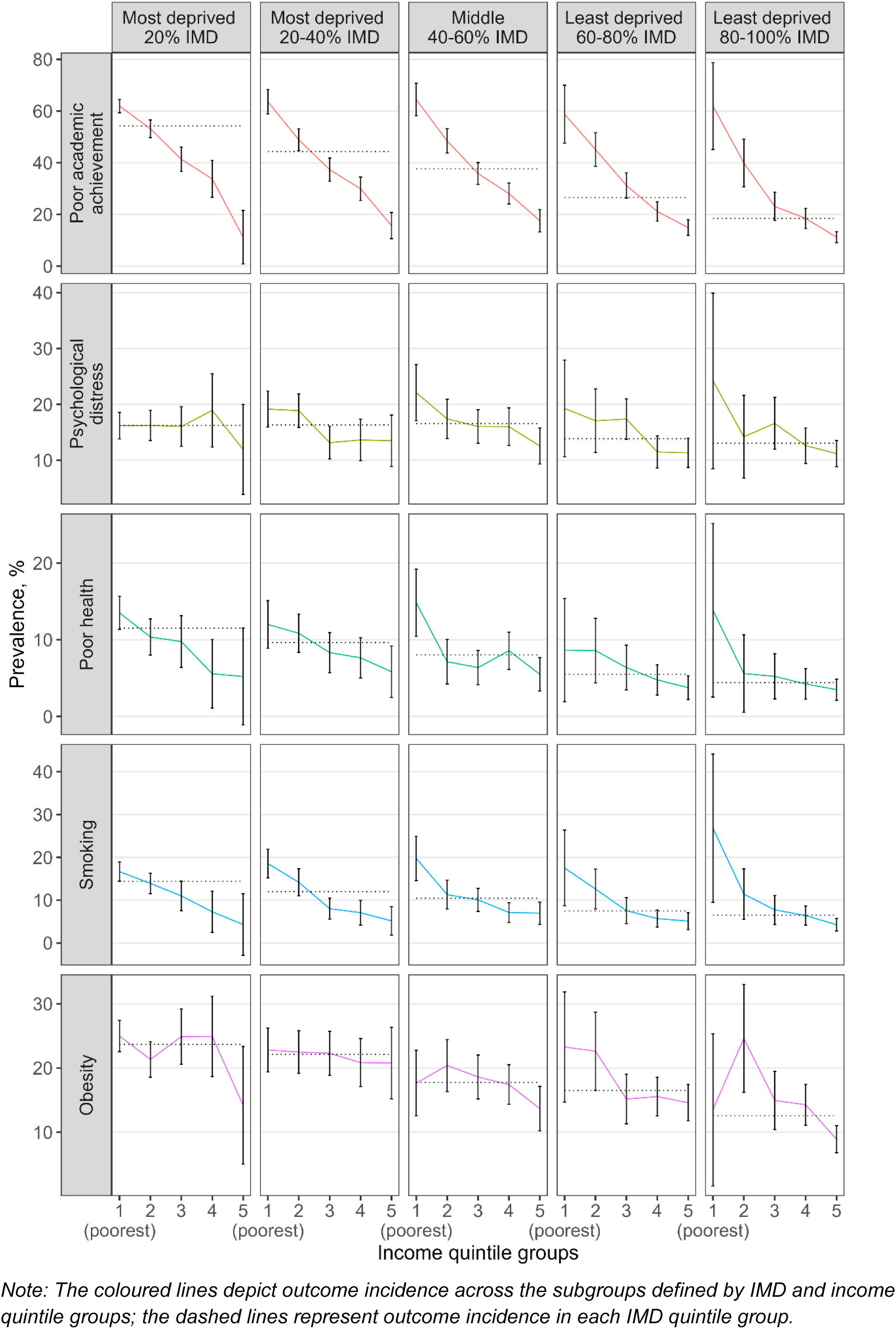
Adverse outcome prevalence at age 17 in each of the IMD-income quintile subgroups

**Figure 2:**
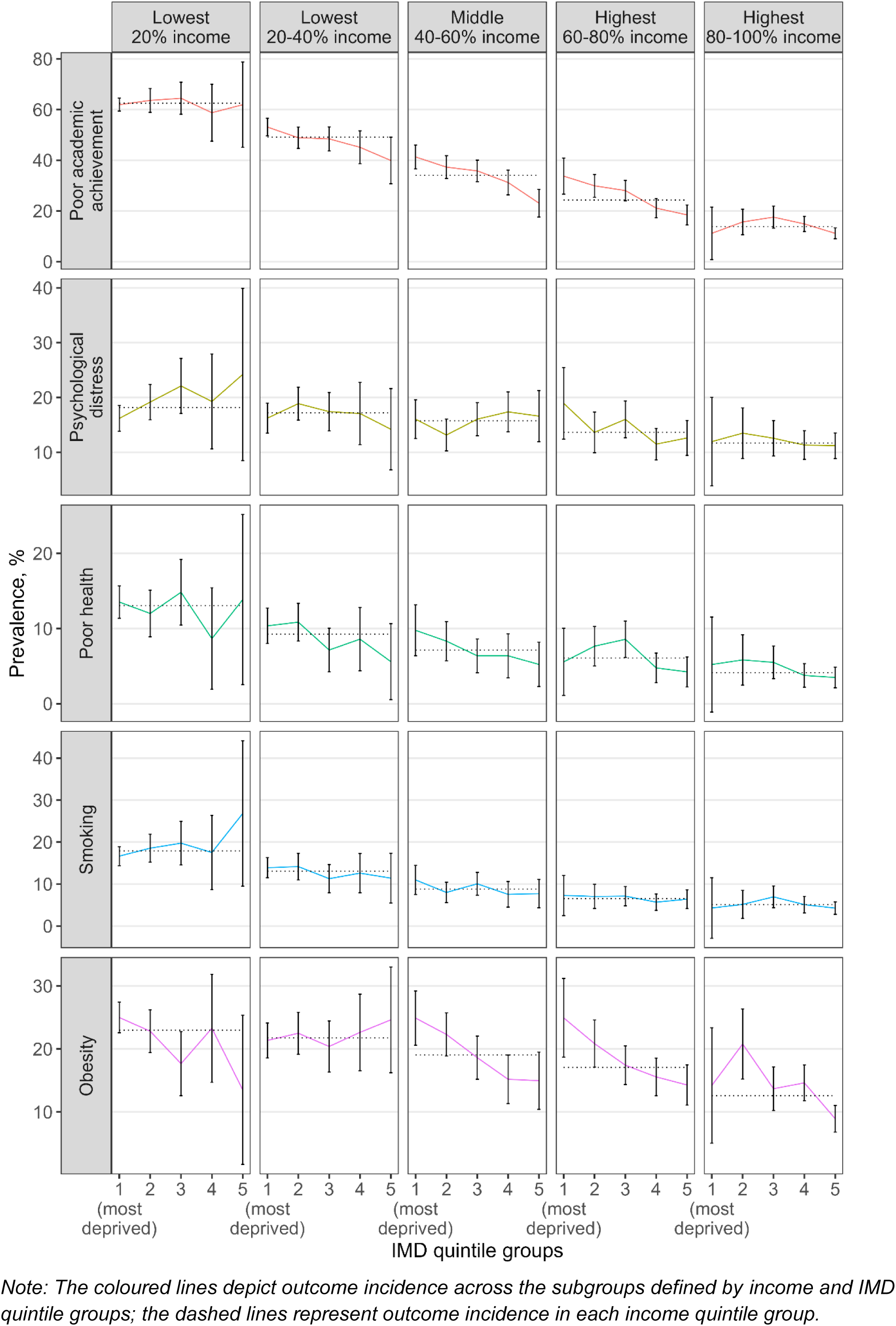
**Adverse outcome prevalence at age 17 in each of the income-IMD quintile subgroups**

For each adverse outcome, we computed the Inequality Gradient Slope Index separately for IMD quintiles, income quintiles, and both combined (appendix Table A3). This index quantifies the percentage point reduction in the likelihood of the adverse outcome when moving from the most deprived to the least deprived child, based on a linear model.^[21]^

Next, we ran modified Poisson regressions to estimate the risk ratios (with 95% confidence intervals) for the incidence of five adverse outcomes at age 17, given the IMD quintiles and income quintiles as predictors in separate models, as well as in a model with both measures together (Table 2). The aim was to compare how well each of the measures predicted the adverse outcomes and the inequality gradient in these outcomes.

**Table 2.** Risk ratios of inequalities in adverse outcomes at age 17 according to quintile of Index of Multiple Deprivation (IMD) or household income, or both.

|  | Model with IMD only |  | Model with income only |  | Model with IMD & income |  |
| --- | --- | --- | --- | --- | --- | --- |
|  | RR | 95% CI | RR | 95% CI | RR | 95% CI |
| **Poor academic achievement** |  |  |  |  |  |  |
| Most deprived 20 % IMD | 2.9*** | 2.6 - 3.3 |  |  | 1.5*** | 1.3 - 1.7 |
| Most deprived 20-40 % IMD | 2.4*** | 2.1 - 2.7 |  |  | 1.5*** | 1.3 - 1.7 |
| Middle 40-60 % IMD | 2.0*** | 1.8 - 2.3 |  |  | 1.5*** | 1.3 - 1.7 |
| Least deprived 60-80 % IMD | 1.4*** | 1.2 - 1.7 |  |  | 1.2** | 1.1 - 1.4 |
| Lowest 20 % income |  |  | 4.5*** | 3.9 - 5.2 | 3.7*** | 3.2 - 4.4 |
| Lowest 20-40 % income |  |  | 3.4*** | 2.9 - 3.9 | 2.9*** | 2.5 - 3.4 |
| Middle 40-60 % income |  |  | 2.3*** | 2.0 - 2.7 | 2.1*** | 1.8 - 2.4 |
| Highest 60-80 % income |  |  | 1.7*** | 1.5 - 2.0 | 1.6*** | 1.4 - 1.9 |
| **Psychological distress** |  |  |  |  |  |  |
| Most deprived 20 % IMD | 1.3* | 1.0 - 1.5 |  |  | 1.0 | 0.8 - 1.3 |
| Most deprived 20-40 % IMD | 1.3* | 1.0 - 1.5 |  |  | 1.1 | 0.9 - 1.3 |
| Middle 40-60 % IMD | 1.3** | 1.1 - 1.5 |  |  | 1.2 | 0.9 - 1.4 |
| Least deprived 60-80 % IMD | 1.1 | 0.9 - 1.3 |  |  | 1.0 | 0.8 - 1.3 |
| Lowest 20 % income |  |  | 1.5*** | 1.3 - 1.8 | 1.5*** | 1.2 - 1.9 |
| Lowest 20-40 % income |  |  | 1.5*** | 1.2 - 1.8 | 1.4*** | 1.2 - 1.7 |
| Middle 40-60 % income |  |  | 1.3** | 1.1 - 1.6 | 1.3** | 1.1 - 1.6 |
| Highest 60-80 % income |  |  | 1.1 | 0.9 - 1.4 | 1.1 | 0.9 - 1.4 |
| **Poor health** |  |  |  |  |  |  |
| Most deprived 20 % IMD | 2.5*** | 1.9 - 3.4 |  |  | 1.6* | 1.1 - 2.2 |
| Most deprived 20-40 % IMD | 2.2*** | 1.6 - 2.9 |  |  | 1.5* | 1.1 - 2.1 |
| Middle 40-60 % IMD | 1.8*** | 1.3 - 2.4 |  |  | 1.4* | 1.0 - 1.9 |
| Least deprived 60-80 % IMD | 1.2 | 0.8 - 1.7 |  |  | 1.1 | 0.7 - 1.6 |
| Lowest 20 % income |  |  | 3.2*** | 2.4 - 4.3 | 2.4*** | 1.7 - 3.5 |
| Lowest 20-40 % income |  |  | 2.1*** | 1.6 - 2.9 | 1.7** | 1.2 - 2.4 |
| Middle 40-60 % income |  |  | 1.8*** | 1.3 - 2.4 | 1.5* | 1.1 - 2.2 |
| Highest 60-80 % income |  |  | 1.5* | 1.1 - 2.1 | 1.4+ | 1.0 - 2.0 |
| **Smoking** |  |  |  |  |  |  |
| Most deprived 20 % IMD | 2.2*** | 1.7 - 2.9 |  |  | 1.1 | 0.8 - 1.5 |
| Most deprived 20-40 % IMD | 1.8*** | 1.4 - 2.4 |  |  | 1.2 | 0.8 - 1.6 |
| Middle 40-60 % IMD | 1.6*** | 1.2 - 2.1 |  |  | 1.2 | 0.9 - 1.6 |
| Least deprived 60-80 % IMD | 1.2 | 0.9 - 1.5 |  |  | 1.0 | 0.7 - 1.4 |
| Lowest 20 % income |  |  | 3.8*** | 2.9 - 5.0 | 3.6*** | 2.5 - 5.0 |
| Lowest 20-40 % income |  |  | 2.5*** | 1.9 - 3.4 | 2.4*** | 1.7 - 3.3 |
| Middle 40-60 % income |  |  | 1.7** | 1.2 - 2.3 | 1.6** | 1.2 - 2.3 |
| Highest 60-80 % income |  |  | 1.4+ | 1.0 - 1.9 | 1.3+ | 1.0 - 1.8 |
| **Obesity** |  |  |  |  |  |  |
| Most deprived 20 % IMD | 1.9*** | 1.6 - 2.2 |  |  | 1.6*** | 1.3 - 1.9 |
| Most deprived 20-40 % IMD | 1.8*** | 1.5 - 2.1 |  |  | 1.5*** | 1.3 - 1.8 |
| Middle 40-60 % IMD | 1.4*** | 1.2 - 1.7 |  |  | 1.3* | 1.0 - 1.5 |
| Least deprived 60-80 % IMD | 1.3** | 1.1 - 1.6 |  |  | 1.3** | 1.1 - 1.6 |
| Lowest 20 % income |  |  | 1.7*** | 1.5 - 2.0 | 1.4*** | 1.2 - 1.7 |
| Lowest 20-40 % income |  |  | 1.7*** | 1.4 - 2.0 | 1.4*** | 1.2 - 1.7 |
| Middle 40-60 % income |  |  | 1.4*** | 1.2 - 1.7 | 1.3** | 1.1 - 1.5 |
| Highest 60-80 % income |  |  | 1.3** | 1.1 - 1.5 | 1.2* | 1.0 - 1.4 |
| Observations | 15,367 |  | 15,367 |  | 15,367 |  |

Note: \*\*\* $p < 0.001$ , \*\* $p < 0.01$ , \* $p < 0.05$ , + $p < 0.10$ ; the least deprived 80-100% IMD and highest 80-100% income are used as a reference. Regression results are based on a modified Poisson model (not adjusted for any confounders). The results from an adjusted regression are in the appendix (Table A4).

In main regression analyses, we did not include any covariates as our aim is to inform government and researchers of the added value of household income in addition to IMD in the context of adverse adolescent health-related outcome prevalence, regardless of other measures available. Adjusting for covariates could potentially alter and likely underestimate any association with IMD or household income.^[22], [23], [24]^ However, in sensitivity analyse, we analysed the extent to which other covariates (child’s sex, eligibility for free school meals, single parent status, maternal age and number of siblings) routinely available in some administrative datasets such as ECHILD or birth registrations, capture the socioeconomic disadvantage reflected by the IMD and household income (appendix Table A4).

Finally, we plotted the receiver operating characteristic (ROC) curves and calculated the area under the curve (AUC) for the Poisson regression models predicting adverse outcomes at age 17, considering the IMD quintile group, the income quintile group, and both predictors together (appendix Figure A4, Figure A5). ROC curves quantify the diagnostic accuracy of prediction models, allowing comparison across multiple models.

The multiple imputation and regression analyses were done using STATA (version 18). The cross tabulation, figures and ROC curve plots were done using R (version R-4.4.0).

## Results

Of the 15,367 adolescents aged 17 years, 7,822 (50.9%) were male and 7,545 (49.1%) were female. The average weekly equivalised household income in nominal prices was £344.3. For around 29% of the sample in early childhood, their IMD quintile coincided with their income quintile. Of those with lowest 20% income, only 52% lived in the lowest IMD quintile, and 23% lived in the middle to richest areas (appendix Figure A3).

Overall, 5,655 (36.8%) of the adolescents achieved poor academic outcomes, 2,351 (15.3%) experienced psychological distress, 1,214 (7.9%) self-reported poor health, 1,583 (10.3%) were regular smokers, and 2,873 (18.7%) were obese (appendix Table A2).

Poor academic achievement was most prevalent in the lowest IMD quintile (54.2%, 95% CI 52.2-56.1; Table 1) and the poorest income quintile (62.6%, 95% CI 60.4-64.7; Table 1). A steep inequality gradient was found with much lower prevalence of poor academic achievement in the least deprived IMD quintile (18.4%, 95% CI 16.4-20.5; Table 1) and richest income quintile (13.9%, 95% CI 12.3-15.4; Table 1).

The prevalence of all adverse outcomes was characterized by apparent inequality gradients in income– i.e., as one moves from the richest to the poorest quintile, the prevalence of adverse outcomes increases (Figure 1). In particular, poor academic achievement, followed by smoking, exhibited the steepest income inequality gradients consistently across all IMD quintile groups. Poor health also showed consistent inequality gradients in income. The income gradients for obesity and psychological distress were more variable, showing moderate gradients within most IMD quintile groups.

Adverse outcomes exhibited moderate to no inequality gradient in neighbourhood IMD within each income quintile (Figure 2). For example, the prevalence of poor academic achievement showed a moderate inequality gradient in IMD across the three middle-income quintiles, indicating that both income and neighbourhood deprivation contribute to poor academic achievement. However, children in the poorest income quintile demonstrated similarly poor attainment regardless of whether they resided in the least or most deprived neighbourhoods (61.9%, 95% CI 59.4-64.5 vs. 61.9%, 95% CI 45.1-78.7; Table 1). Those in the highest income quintile experienced the lowest rates of poor attainment in all IMD groups, with minimal variation within the high-income quintile according to neighbourhood deprivation levels.

The inequality gradients in IMD for psychological distress and poor health varied from weak to no gradient, particularly among those with the lowest 20% income. However, for obesity, neighbourhood IMD was associated with a moderate gradient of declining obesity among children in the three highest quintiles of household income who live in less deprived neighbourhoods. Smoking appears to relate to low income rather than neighbourhood quintile.

The slope index of inequality was higher across income quintiles than across IMD quintiles (appendix Table A3). For example, based on this index, we estimated a 61.1 percentage point reduction in the probability of poor academic achievement when moving from the poorest to the richest child in terms of household income; and a 44.1 percentage point reduction when moving from the most deprived to the least deprived child according to IMD. Similarly, we estimated an 8.3 percentage point reduction in the probability of psychological distress when moving from the poorest to the richest child; and a 4.4 percentage point reduction when moving from the most deprived to the least deprived child.

The regression analyses indicated that income was a stronger predictor of adverse outcomes than IMD, for all outcomes except for obesity (Table 2). The coefficients for income were higher than for IMD in models using each exposure alone and in the combined model. Coefficients for IMD were no longer significant at the 5% level after including income when predicting psychological distress and smoking, and were reduced, but still significant when predicting poor educational attainment, poor health and obesity. Income and IMD had effects of a similar magnitude for obesity, and IMD had a stronger effect in the combined model. These findings suggest independent effects of household income and neighbourhood IMD on poor educational attainment, poor health and obesity, reflecting the added value of including both measures. All models that included income consistently predicted more frequent adverse outcomes in the poorest subgroups.

The finding of income being a stronger predictor than IMD for all outcomes except for obesity was supported by our sensitivity analyses.

First, the ROC curve analysis (appendix Figure A4) shows that the combined model (IMD & income) consistently returns the highest area under curve (AUC) when predicting the incidence of all five adverse outcomes, so both income and IMD have the highest predictive power when used together. However, income is the most important predictor: moving from a model with IMD quintiles as the sole predictors to a model with income quintiles as the predictors provides the highest increase in AUC (statistically significant at 95% level)^[25]^, particularly, when predicting the incidence of poor academic achievement, poor health, and smoking. This implies that using income instead of IMD as a predictor for these outcomes substantially increases the sensitivity while maintaining the specificity of the model.

Our regression analyses adjusted for the child’s sex and socioeconomic characteristics (appendix Table A4), and the corresponding ROC curves (appendix Figure A4), found that household income during early childhood remained a relatively stronger predictor of adverse outcomes at age 17 and inequality in these outcomes.

## Discussion

### Main findings

Household income in early childhood was a stronger and more consistent predictor than neighbourhood IMD for four of the five health-related outcomes at age 17. IMD was a slightly stronger predictor for obesity. Adverse outcomes increased consistently with decreasing household income, both in analyses based on income alone and in combination with IMD. In contrast, neighbourhood IMD was less consistently associated with adverse outcomes once income was considered.

Including both income and IMD in the analysis revealed patterns that would otherwise go unnoticed. Neighbourhood IMD was not associated with smoking or psychological distress in any of the income quintiles, nor with educational attainment in the poorest and richest income quintiles. Using IMD alone would miss the steep gradients of poorer educational attainment and increased smoking with decreasing household income within each IMD quintile.

### What is already known on this topic?

Given the lack of UK evidence highlighted in a recent systematic review^[15]^, it is not known which is a better proxy for early-years disadvantage: IMD or household income or both.

IMD and household income are different measures, leading to variations in their associations with adverse adolescent outcomes. The income deprivation component of IMD measures the proportion of low-income households meeting benefit thresholds, while household income is recorded for each child and grouped into quintiles across the income distribution (see appendix). IMD also captures multiple deprivation indicators, including low income, unemployment, and neighbourhood measures of education, health, crime, and access to amenities (listed in Table A1). Neighbourhood IMD is less sensitive and specific for detecting income-deprived households^[12–14]^, resulting in weaker inequality gradients compared to household-level measures. However, the multi-dimensionality of IMD may capture neighbourhood effects on young families regardless of income.^[26]^

### What this study adds

We found that household income in early childhood is a stronger predictor of adverse health outcomes in adolescence than neighbourhood IMD. However, combining income and IMD adds useful information for understanding subgroups at highest risk. Adding household income groupings to IMD could improve understanding of policies that affect household income, expose larger disparities in outcomes than is evident using IMD alone, and improve targeting of families who stand to benefit from early years interventions. Our findings suggest that the current core20plus5 approach advocated by NHS England ^[11]^, is a poor predictor of children most at risk and does not consider how to address the gradient of disadvantage through policies that affect household income.^[27]^

Our findings strengthen arguments to widen the use and development of measure of individual or household income to address what has been called the inverse evidence law, whereby there is least evidence on the upstream determinants of health inequalities that are likely to have most impact on health but are most difficult to research.^[10]^ Research on health inequalities from the Nordic countries, the Netherlands and North America, reports use of income alongside area-based deprivation indices.^[26, 28–35]^ A recent US report describes a comprehensive income dataset that incorporates nearly all taxable income which is used to address under-reporting in surveys and for tax administration, forecasting and research.^[28, 29]^

In the UK, improving information on costs and income is a priority for population studies on health and deprivation, as identified in a review and poverty experts.^[36]^ Currently, the Office of National Statistics (ONS) derives household income measures, but allows their use only at the area level.^[37, 38]^ However, recent ONS research, using employment data linked to health records, could serve as an exemplar for broader use of anonymized income data linked to administrative healthcare data for research.^[39]^

### Limitations of this study

First, the study sample was not large enough to draw statistical conclusions about the differences between the 25 subgroups combining household income and neighbourhood IMD. Second, missing data disproportionately affected disadvantaged groups but was addressed by multiple imputation^[18, 19]^, and by averaging income over three rounds of interviews in the early years. Third, we used parent-reported income as administrative data on household income is not yet available for England: differences between these sources need to be evaluated. Fourth, we used neighbourhood IMD, which includes population health outcomes, rather than restrict to the income deprivation domain of IMD. This was because IMD is a tool for policy and public health and restriction to income deprivation makes very little difference.^[40]^

## Conclusion

Household income in early childhood was a stronger predictor of health-related outcomes at age 17 than neighbourhood deprivation and was associated with steeper inequality gradients than currently recognised. Public health intervention in early childhood could be more effective if guided by household income as well as neighbourhood deprivation.

## Data Availability

The datasets analysed during the current study are available through the UK Data Service (https://ukdataservice.ac.uk/). For further information on the Millennium Cohort Study, including data documentation and metadata, please visit the Centre for Longitudinal Studies website (https://cls.ucl.ac.uk/cls-studies/millennium-cohort-study/).

## Acknowledgements

We would like to thank our colleagues from the Children and Families Policy Research Unit, the Department for Health and Social Care, and the University of York for their useful discussions and feedback during the presentation of this work. We also express our gratitude to Aase Villadsen, who led the analyses of the cohort and variables for a study previously published in The Lancet, which we re-used and adapted for this study. Finally, we are grateful to the Centre for Longitudinal Studies (CLS), UCL Social Research Institute, for the use of the MCS data, and to the UK Data Service for making these data available. However, neither CLS nor the UK Data Service bear any responsibility for the analysis or interpretation of these data.

## Data availability

The datasets analysed during the current study are available through the UK Data Service <u>here</u>. For further information on the Millennium Cohort Study, including data documentation and metadata, please visit the Centre for Longitudinal Studies <u>website</u>.

