## Appendix for "Does household income predict health and educational outcomes in childhood better than neighbourhood deprivation?"

### A1. Multiple imputations and weighting

Multiple imputation with chained equations was used to address missing data, which was primarily due to attrition over time and low response rates to specific MCS survey questions. We imputed data back to the age 3 survey sample (sweep 2), chosen as our primary sweep for imputation since it includes the 1,389 additional families missed in sweep 1 (see Figure A1 below). [1, 2] We created 30 imputed datasets. To enhance the accuracy of our estimates, several auxiliary variables from various MCS sweeps predictive of the five outcomes at age 17 were used in the imputation process. These variables included data on cognitive assessments, child mental health, physical health, substance use, and obesity. [3] After imputation, the final analysis sample comprised 15,367 cohort members. All analyses were conducted after applying weights provided in the MCS dataset to adjust for attrition between the initial survey and the age 5 survey, as well as for the complex sampling design.

1. Mostafa T, Narayanan M, Pongiglione B, et al. Missing at random assumption made more plausible: evidence from the 1958
British birth cohort. Journal of Clinical Epidemiology 2021;136:44-54

2. Mishra S, Khare D. On comparative performance of multiple imputation methods for moderate to large proportions of missing
data in clinical trials: a simulation study. Med Stat Inform 2014;2(1):9

3. Von Hippel P, Lynch J. Efficiency gains from using auxiliary variables in imputation. arXiv 2013;1311.5249

**Figure A1. Deriving the primary analysis dataset using MCS data**

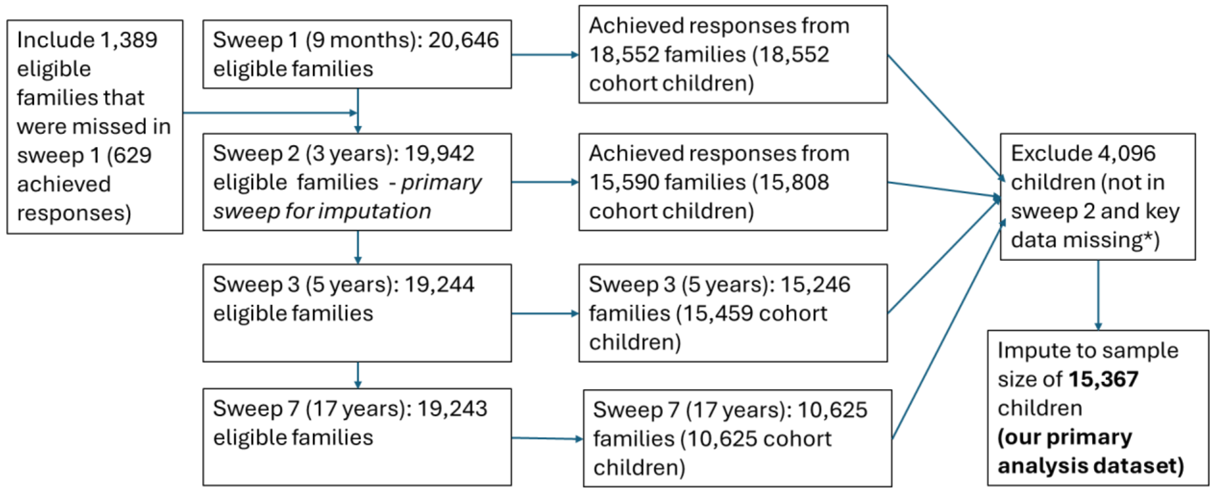


*Note: *Before imputing we exclude the 3,655 children not in the primary sweep 2. We also exclude 441 other children with key missing data (ethnicity and month of birth) and all twins and triplets due to low numbers and because these are likely to cause problems with multiple imputation due to collinearity.*

**Table A1. Definitions of the Socioeconomic Measures and Adverse Outcomes**

| **Measure** | **Description** |
|---|---|
| **Socioeconomic measures during early childhood** | |
| Household-level income quintile groups | Quintile groups based on the MCS early childhood household income measure at 9 months, 3 and 5 years. MCS income measure is based on banded self-reported household income (after tax and other deductions but before housing costs) reported by the main caregiver at ages 9 months, 3 years and 5 years, collected by a home interviewer. Banded responses were used to impute the continuous income measure, which was equivalised using OECD modified scales, then averaged over the three sweeps and ranked to calculate the income quintile groups. |
| Lower super output area (LSOA)-level Index of Multiple Deprivation (IMD) quintile groups | Quintile groups based on the IMD 2004, geographically linked to the MCS households at the LSOA-level at each sweep (9 months, 3 and 5 years), ranked separately for England, Scotland, Wales and Northern Ireland. The deciles at 9 months, 3 years, 5 years were averaged over the three sweeps and then rounded. Quintile group values were calculated by combining decile values.<br><br>IMD measures relative deprivation across England at the small area level averaging 650 households and 1,600 individuals (called lower super output area or LSOA (ref tech report). Income deprivation is a central component of IMD. In the current IMD (2019) the proportion of children or adults living in families in receipt of benefits or whose equivalised income (excluding housing benefits) is below 60 per cent of the national median before housing costs (p29 technical report). Income deprivation accounts for 22.5% of the IMD score.<br><br>Employment deprivation accounts for a further 22.5% of IMD, and indicates working age adults not in employment, denoted by out of work benefits. For the 2019 IMD, both income deprivation and employment were based on 2015 data from the HMRC or data from the Dept of Work and Pensions (DWP)).<br><br>Five other domains account for 55% of the IMD score and include:<br>a) education and skills (average school attainment of children and proportion of adults without skills in the LSOA; 13.5% of IMD score)<br>b) a composite health measure using: years of potential life lost (based on deaths), benefit claims for disability (DWP), emergency admissions or prescribing related to mental health, and suicide (at LSOA level, accounting for 13.5% of IMD score);<br>c) recorded crime events (with weighted subcategories) per LSOA, provided by National Police Chiefs Council and the Home Office (9.3% of IMD score); |

| | |
|---|---|
| | d) barriers to housing and services based on distance to amenities, overcrowding, homelessness and housing affordability (9.3% of IMD score);<br>e) the living environment deprivation domain based on housing disrepair/modernisation, air quality and road traffic accidents (9.3% of IMD score).<br><br>Prior updates were 2004,2007, 2010 and 2015. We used the 2004 for this study as closest to the early years of study participants of MCS born in 2001--02. |
| Number of siblings | Number of siblings recorded as present in household at age 9 months, grouped into the following categories: none, 1, 2, 3, 4 or more. Data has been self-reported by the main parent/carer in a household interview. |
| Maternal age | Age of the natural mother when child is 9 months old, grouped into the following categories: "under 16", "16 to 19", "20 to 24", "25 to 29", "30 to 34", "35 to 39", "40 or more". Based on self-reported data by the main parent/carer in a household interview. |
| Free school meals | Indicator of child eligible for free school meals at age 5. Based on self-reported data by the main parent/carer in a household interview. |
| Single parent status | Indicator of single parent/carer status at age 9 months of the child. Based on self-reported data by the main parent/carer in a household interview |
| **Adverse outcomes at age 17** | |
| Poor academic achievement | Indicator of not achieving five or more GCSEs (including in maths and English) graded C or above, or five or more N5s (including in maths<br>and English) graded D or above. Based on self-reported exam results<br>at the end of secondary school. |
| Psychological distress | Indicator of psychological distress, based on self-reported six item Kessler scale (range 0-24) exceeding clinically determined threshold (≥13) (Kessler et al, 2003). |
| Poor health | Indicator of poor health based on self-rated health, assessed by selecting an answer to the question "How would you describe your health generally?" (with response options: excellent/very good/good/fair/poor). Responses of "fair" and "poor" were classified as being in poor health. |
| Smoking | Self-reported regular smoking of more than six cigarettes per week (excluding e-cigarettes). |
| Obesity | Indicator of obesity based on the adolescents' weight and height, taken by the interviewer at the participant's home, and classified using the obesity threshold from the British 1990 (UK90) growth reference chart for children (Freeman et al. 1995). |

**Table A2. Prevalence of baseline characteristics before age five and outcomes at age 17 for the imputed and source study population (non-imputed)**

| Outcomes and characteristics | Primary analysis dataset: imputed to age 3 years sample (*N=15,367*) | Sensitivity analysis dataset: non-imputed age 17 sample (*N=7,764*) |
|---|---|---|
| **Sex** | | |
| Female | 49.1% [48.2-50.0] | 49.1% [47.1-51.0] |
| Male | 50.9% [50.0-51.8] | 51.0% [49.0-52.9] |
| **Ethnicity** | | |
| White | 86.5% [85.9-87.0] | 86.2% [85.0-87.4] |
| Mixed | 3.2% [2.9-3.5] | 3.6% [2.9-4.3] |
| Indian | 1.9% [1.6-2.1] | 1.7% [1.4-2.1] |
| Pakistani and Bangladeshi | 4.3% [4.0-6.6] | 3.8% [3.4-4.3] |
| Black and Black British | 2.8% [2.5-3.1] | 3.2% [2.6-4.0] |
| Other ethnic group | 1.3% [1.1-1.5] | 1.4% [1.0-2.1] |
| **Country** | | |
| England | 82.7% [82.1-83.2] | 84.5% [83.4-85.5] |
| Wales | 4.9% [4.7-5.1] | 5.1% [4.6-5.7] |
| Scotland | 8.8% [8.4 -9.3] | 7.3%[6.7-8.1] |
| Northern Ireland | 3.6% [3.4-3.8] | 3.1% [2.7-3.5] |
| **Household-level income (averaged 9 months-5 years)** | | |
| Self-reported weekly income, £ | 344.3 [340.5-348.1] | 330.3 [327.0-333.6] |
| Lowest 20% income | 17.7% [17.0-18.4] | 14.2% [13.1-15.3] |
| Lowest 20-40% income | 18.5% [17.8- 19.3] | 16.3% [15.3-17.4] |
| Middle 40-60% income | 20.3% [19.6-21.1] | 20.9% [19.8-22.0] |
| Highest 60-80% income | 20.9% [20.2-21.7] | 23.0% [21.9-24.2] |
| Highest 80-100% income | 22.6% [21.8-23.4] | 25.7% [24.5-26.9] |
| **LSOA-level Index of Multiple Deprivation (IMD) (averaged 9 months-5 years)** | | |
| Most deprived 20% IMD | 20.8% [20.1-21.6] | 15.7% [14.8-16.7] |
| Most deprived 20-40% IMD | 20.0% [19.3-20.7] | 18.3% [17.3-19.3] |
| Middle 40-60% IMD | 21.5% [20.7-22.3] | 22.7% [21.5-23.9] |
| Least deprived 60-80% IMD | 19.7% [18.9-20.5] | 22.5% [21.3-23.6] |
| Least deprived 80-100% IMD | 18.0% [17.2-18.7] | 20.8% [19.8-21.9] |
| **Number of siblings (9 months)** | | |
| None | 41.4% [40.5-42.4] | 39.7 [38.8-40.6] |
| 1 | 36.3% [35.3-37.2] | 34.5 [33.6-35.4] |
| 2 | 15.2% [14.5-15.9] | 14.1 [13.5-14.7] |
| 3 | 5.0% [4.6-5.4] | 4.6 [4.3-5.0] |
| 4+ | 2.1% [1.8-2.3] | 7.1 [6.6-7.6] |
| **Maternal age (9 months)** | | |
| Under 16 | 0.3% [0.2-0.4] | 0.3 [0.2-0.4] |
| 16 to 19 | 7.7% [7.2-8.2] | 7.6 [7.1-8.1] |
| 20 to 24 | 16.9% [16.2-17.5] | 16.5 [16.8-17.2] |
| 25 to 29 | 27.8% [27.0-28.6] | 27.2 [26.4-28.1] |

| | | |
|---|---|---|
| 30 to 34 | 30.3% [29.4-31.1] | 29.9 [29.0-30.7] |
| 35 to 39 | 14.8% [14.1-15.4] | 14.6 [13.9-15.2] |
| 40 or over | 2.3% [2.0-2.5] | 4.0 [3.6-4.4] |
| **Other socioeconomic markers** | | |
| Received free school meals (age 5) | 13.9% [13.3-14.6] | 13.1 [12.5-13.8] |
| Single parent status (9 months) | 14.9% [14.2-15.6] | 14.5 [13.8-15.1] |
| **Individual adverse outcomes (age 17)** | | |
| Poor academic achievement | 36.8% [35.8-37.8] | 31.4% [30.3-32.6] |
| Psychological distress | 15.3% [14.6-16.0] | 16.1% [15.2-16.9] |
| Poor health | 7.9% [7.3-8.5] | 7.1% [6.5-7.6] |
| Smoking | 10.3% [9.7-10.9] | 8.3% [7.6-8.9] |
| Obesity | 18.7% [17.9-19.5] | 20.3% [19.2-21.2] |

*Note: The figures are adjusted for the complex sampling design and for attrition between the initial survey and the age 5 survey used as the imputation samples. The weighting means that the sample is representative of children born in the UK at the start of the millennium. Self-reported income is weekly OECD equivalised income after tax and other deductions but before housing costs.*

**Figure A2. Sample size in each income-IMD subgroup across the three datasets**

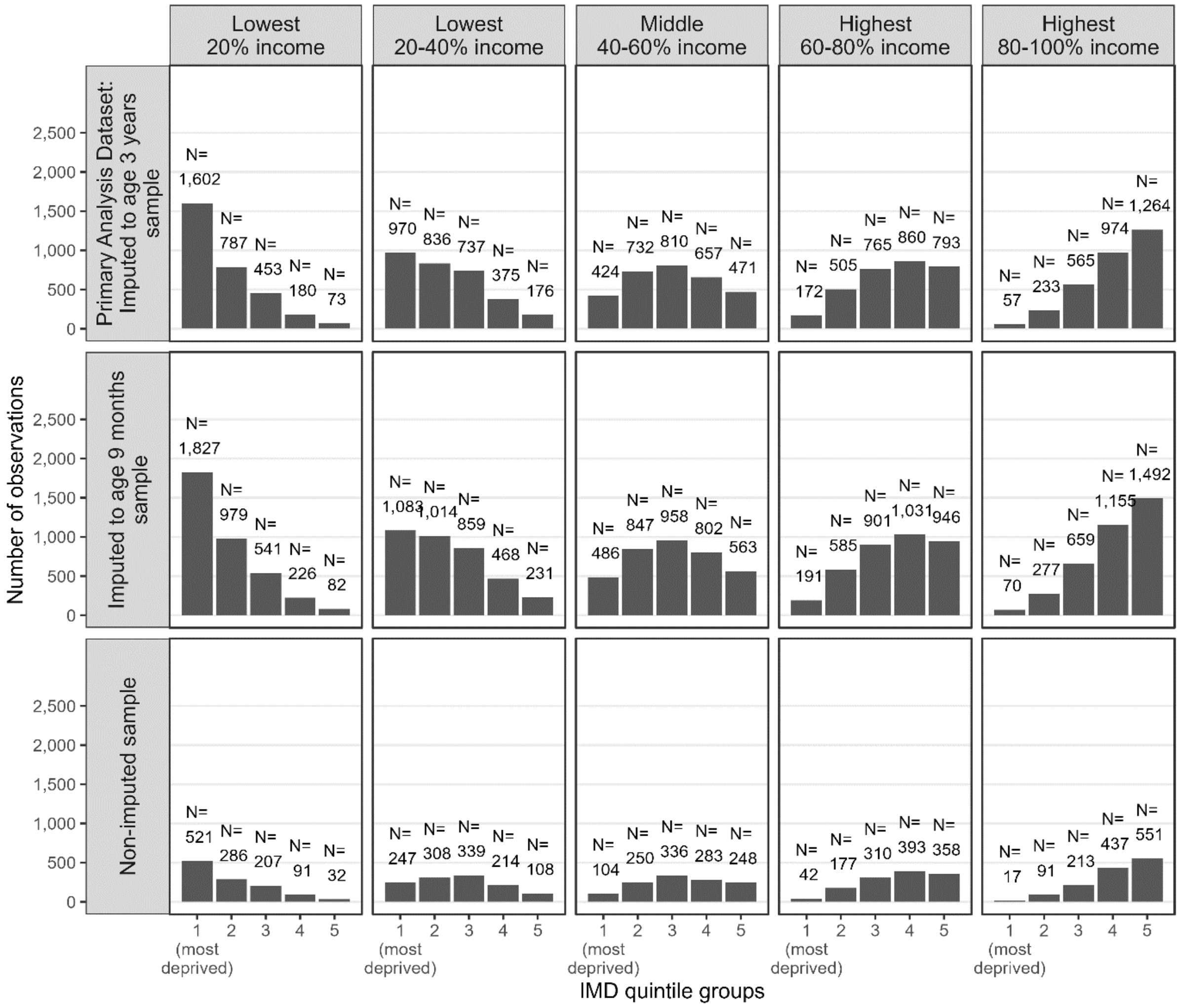


*Note: The values represent the number of children in each subgroup defined by IMD quintile and income quintile groups in early childhood. The figures are adjusted for the complex sampling design and for attrition between the initial survey and the age 5 survey used as the imputation samples. The weighting means that the sample is representative of children born in the UK at the start of the millennium.*

**Figure A3: Distribution of individuals across the IMD-income subgroups in the primary analysis dataset**

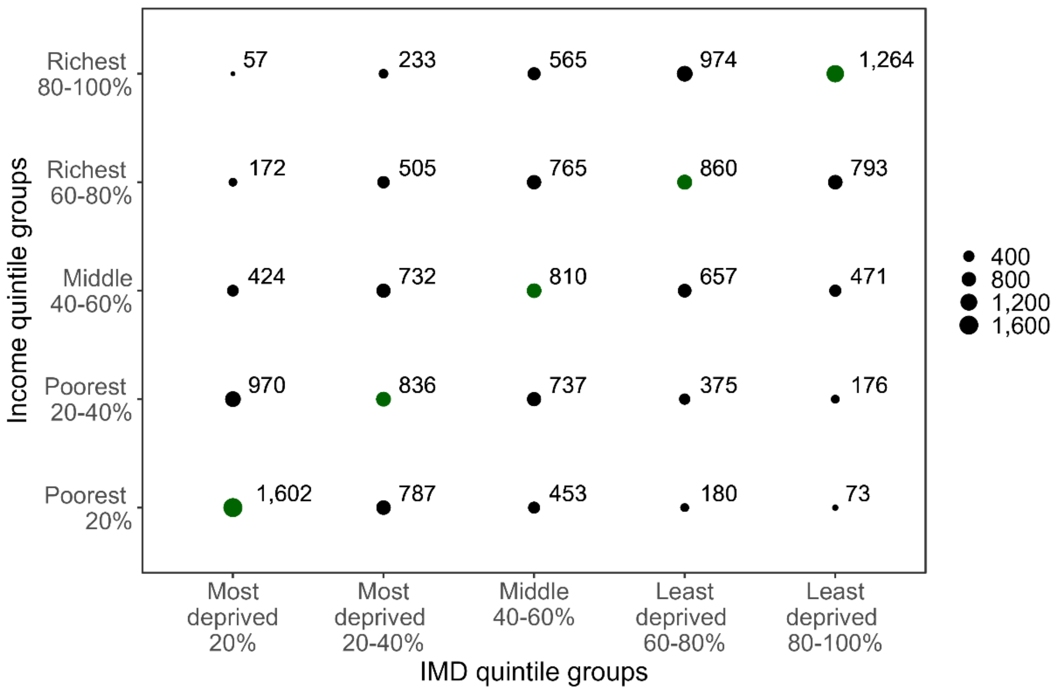


*Note: The values represent the number of children in the primary analysis dataset in each subgroup defined by IMD quintile and income quintile groups in early childhood. The values on the diagonal line marked green represent The figures are adjusted for the complex sampling design and for attrition between the initial survey and the age 5 survey used as the imputation samples. The weighting means that the sample is representative of children born in the UK at the start of the millennium.*

**Table A3. Slope index of inequality across subgroups with different measures of disadvantage**

| Adverse outcomes at age 17 | **IMD only** (5 subgroups) | **Income only** (5 subgroups) | **IMD and income** (25 subgroups) |
|---|---|---|---|
| | | **Slope index** | |
| Poor academic achievement | -44.1 | -61.1 | -65.4 |
| Psychological distress | -4.4 | -8.3 | -9.9 |
| Poor health | -9.1 | -10.5 | -10.6 |
| Smoking | -10.0 | -16.0 | -19.0 |
| Obesity | -13.7 | -12.8 | -10.6 |

Note: Percentage point reduction in the probability of the adverse outcome if you move from the poorest child to the richest child based on a linear model.

**Table A4. Risk ratios with confidence intervals of inequalities in adverse outcomes at age 17 (adjusted for control variables *in italics*)**

| | Model with IMD only | | Model with income only | | Model with IMD & income | |
|---|---|---|---|---|---|---|
| | RR | 95% CI | RR | 95% CI | RR | 95% CI |
| **Poor academic achievement** | | | | | | |
| Most deprived 20 % IMD | 1.8*** | 1.6 - 2.1 | | | 1.4*** | 1.2 - 1.6 |
| Most deprived 20-40 % IMD | 1.8*** | 1.6 - 2.0 | | | 1.4*** | 1.2 - 1.6 |
| Middle 40-60 % IMD | 1.7*** | 1.5 - 2.0 | | | 1.4*** | 1.2 - 1.6 |
| Least deprived 60-80 % IMD | 1.3*** | 1.1 - 1.5 | | | 1.2* | 1.0 - 1.4 |
| Lowest 20 % income | | | 3.1*** | 2.7 - 3.6 | 2.7*** | 2.3 - 3.2 |
| Lowest 20-40 % income | | | 2.7*** | 2.3 - 3.1 | 2.4*** | 2.0 - 2.8 |
| Middle 40-60 % income | | | 2.0*** | 1.7 - 2.3 | 1.8*** | 1.6 - 2.1 |
| Highest 60-80 % income | | | 1.6*** | 1.4 - 1.9 | 1.5*** | 1.3 - 1.8 |
| *Male sex* | 1.3*** | 1.2 - 1.3 | 1.3*** | 1.2 - 1.4 | 1.3*** | 1.2 - 1.4 |
| *Child received free school meals at age 5* | 1.3*** | 1.2 - 1.4 | 1.2*** | 1.1 - 1.3 | 1.2*** | 1.1 - 1.2 |
| *Single parent status at birth* | 1.3*** | 1.2 - 1.4 | 1.1* | 1.0 - 1.2 | 1.1* | 1.0 - 1.2 |
| *Maternal age: under 16* | 1.3 | 0.7 - 2.3 | 1.0 | 0.6 - 1.9 | 1.0 | 0.6 - 1.8 |
| *Maternal age: 16 to 19* | 1.9*** | 1.7 - 2.2 | 1.5*** | 1.3 - 1.7 | 1.5*** | 1.3 - 1.7 |
| *Maternal age: 20 to 24* | 1.6*** | 1.5 - 1.8 | 1.3*** | 1.2 - 1.5 | 1.3*** | 1.2 - 1.4 |
| *Maternal age: 25 to 29* | 1.3*** | 1.2 - 1.4 | 1.2*** | 1.1 - 1.3 | 1.2*** | 1.1 - 1.3 |
| *Maternal age: 35 to 39* | 0.9 | 0.8 - 1.0 | 1.0 | 0.9 - 1.1 | 1.0 | 0.9 - 1.1 |
| *Maternal age: 40 or over* | 0.9 | 0.8 - 1.2 | 1.0 | 0.8 - 1.2 | 1.0 | 0.8 - 1.2 |
| *Number of siblings: 1* | 1.2*** | 1.1 - 1.3 | 1.1** | 1.0 - 1.2 | 1.1** | 1.0 - 1.2 |
| *Number of siblings 2* | 1.6*** | 1.4 - 1.7 | 1.3*** | 1.2 - 1.4 | 1.3*** | 1.2 - 1.4 |
| *Number of siblings 3* | 1.8*** | 1.6 - 2.0 | 1.4*** | 1.2 - 1.6 | 1.4*** | 1.2 - 1.6 |
| *Number of siblings 4 or more* | 2.2*** | 1.9 - 2.5 | 1.6*** | 1.3 - 1.8 | 1.5*** | 1.3 - 1.8 |
| **Psychological distress** | | | | | | |
| Most deprived 20 % IMD | 1.1 | 0.9 - 1.4 | | | 1.0 | 0.8 - 1.2 |
| Most deprived 20-40 % IMD | 1.1 | 0.9 - 1.4 | | | 1.0 | 0.9 - 1.3 |
| Middle 40-60 % IMD | 1.2* | 1.0 - 1.4 | | | 1.1 | 0.9 - 1.4 |
| Least deprived 60-80 % IMD | 1.0 | 0.9 - 1.2 | | | 1.0 | 0.8 - 1.2 |
| Lowest 20 % income | | | 1.5*** | 1.2 - 1.9 | 1.5*** | 1.2 - 2.0 |
| Lowest 20-40 % income | | | 1.5*** | 1.2 - 1.8 | 1.4*** | 1.2 - 1.8 |
| Middle 40-60 % income | | | 1.4** | 1.1 - 1.6 | 1.3** | 1.1 - 1.6 |
| Highest 60-80 % income | | | 1.2 | 1.0 - 1.4 | 1.1 | 0.9 - 1.4 |
| *Male sex* | 0.5*** | 0.4 - 0.5 | 0.5*** | 0.4 - 0.5 | 0.5*** | 0.4 - 0.5 |
| *Child received free school meals at age 5* | 1.1 | 0.9 - 1.3 | 1.0 | 0.8 - 1.2 | 1.0 | 0.9 - 1.2 |
| *Single parent status at birth* | 1.1 | 0.9 - 1.3 | 1.0 | 0.8 - 1.2 | 1.0 | 0.8 - 1.2 |
| *Maternal age: under 16* | 0.6 | 0.1 - 2.5 | 0.5 | 0.1 - 2.3 | 0.5 | 0.1 - 2.3 |
| *Maternal age: 16 to 19* | 1.2+ | 1.0 - 1.6 | 1.1 | 0.8 - 1.4 | 1.1 | 0.8 - 1.4 |
| *Maternal age: 20 to 24* | 1.2+ | 1.0 - 1.4 | 1.1 | 0.9 - 1.3 | 1.1 | 0.9 - 1.3 |
| *Maternal age: 25 to 29* | 1.1 | 0.9 - 1.2 | 1.0 | 0.9 - 1.2 | 1.0 | 0.9 - 1.2 |
| *Maternal age: 35 to 39* | 1.0 | 0.9 - 1.2 | 1.1 | 0.9 - 1.3 | 1.1 | 0.9 - 1.3 |
| *Maternal age: under 16* | 1.1 | 0.7 - 1.5 | 1.0 | 0.7 - 1.5 | 1.0 | 0.7 - 1.5 |
| *Number of siblings: 1* | 1.1 | 0.9 - 1.2 | 1.0 | 0.9 - 1.1 | 1.0 | 0.9 - 1.2 |
| *Number of siblings 2* | 1.1 | 0.9 - 1.3 | 1.0 | 0.8 - 1.2 | 1.0 | 0.8 - 1.2 |
| *Number of siblings 3* | 1.1 | 0.9 - 1.4 | 1.0 | 0.7 - 1.3 | 1.0 | 0.8 - 1.3 |
| *Number of siblings 4 or more* | 1.2 | 0.8 - 1.7 | 1.0 | 0.7 - 1.5 | 1.0 | 0.7 - 1.5 |
| **Poor health** | | | | | | |
| Most deprived 20 % IMD | 1.9*** | 1.3 - 2.8 | | | 1.5* | 1.0 - 2.3 |
| Most deprived 20-40 % IMD | 1.8*** | 1.3 - 2.5 | | | 1.5* | 1.0 - 2.2 |
| Middle 40-60 % IMD | 1.6** | 1.2 - 2.3 | | | 1.4+ | 1.0 - 2.0 |

| | | | | | | |
|---|---|---|---|---|---|---|
| Least deprived 60-80 % IMD | 1.2 | 0.8 - 1.7 | | | 1.0 | 0.7 - 1.5 |
| Lowest 20 % income | | | 2.6*** | 1.8 - 3.8 | 2.1*** | 1.4 - 3.2 |
| Lowest 20-40 % income | | | 1.9*** | 1.4 - 2.8 | 1.6* | 1.1 - 2.4 |
| Middle 40-60 % income | | | 1.6** | 1.1 - 2.3 | 1.4+ | 1.0 - 2.1 |
| Highest 60-80 % income | | | 1.5* | 1.0 - 2.1 | 1.4+ | 1.0 - 2.0 |
| *Male sex* | 0.8* | 0.7 - 1.0 | 0.8* | 0.7 - 1.0 | 0.8* | 0.7 - 1.0 |
| *Child received free school meals at age 5* | 1.4** | 1.1 - 1.8 | 1.2 | 1.0 - 1.6 | 1.2 | 0.9 - 1.6 |
| *Single parent status at birth* | 1.0 | 0.8 - 1.4 | 0.9 | 0.7 - 1.2 | 0.9 | 0.7 - 1.2 |
| *Maternal age: under 16* | 1.4 | 0.4 - 5.3 | 1.2 | 0.3 - 5.1 | 1.1 | 0.3 - 5.0 |
| *Maternal age: 16 to 19* | 1.5+ | 1.0 - 2.1 | 1.2 | 0.8 - 1.9 | 1.2 | 0.8 - 1.8 |
| *Maternal age: 20 to 24* | 1.3+ | 1.0 - 1.7 | 1.2 | 0.9 - 1.6 | 1.2 | 0.9 - 1.5 |
| *Maternal age: 25 to 29* | 1.2 | 0.9 - 1.5 | 1.2 | 0.9 - 1.5 | 1.1 | 0.9 - 1.5 |
| *Maternal age: 35 to 39* | 1.1 | 0.8 - 1.4 | 1.1 | 0.8 - 1.4 | 1.1 | 0.8 - 1.4 |
| *Maternal age: under 16* | 0.9 | 0.5 - 1.7 | 0.7 | 0.4 - 1.5 | 0.7 | 0.4 - 1.5 |
| *Number of siblings: 1* | 1.0 | 0.8 - 1.3 | 1.0 | 0.8 - 1.2 | 1.0 | 0.8 - 1.2 |
| *Number of siblings 2* | 1.3* | 1.0 - 1.7 | 1.2 | 0.9 - 1.5 | 1.2 | 0.9 - 1.6 |
| *Number of siblings 3* | 1.3 | 0.9 - 1.8 | 1.1 | 0.7 - 1.6 | 1.1 | 0.7 - 1.6 |
| *Number of siblings 4 or more* | 1.5 | 0.9 - 2.4 | 1.2 | 0.7 - 2.1 | 1.2 | 0.7 - 2.1 |
| | | | **Smoking** | | | |
| Most deprived 20 % IMD | 1.1 | 0.8 - 1.4 | | | 0.9 | 0.7 - 1.3 |
| Most deprived 20-40 % IMD | 1.2 | 0.9 - 1.5 | | | 1.0 | 0.8 - 1.4 |
| Middle 40-60 % IMD | 1.2 | 0.9 - 1.6 | | | 1.1 | 0.8 - 1.5 |
| Least deprived 60-80 % IMD | 1.1 | 0.8 - 1.4 | | | 1.0 | 0.8 - 1.4 |
| Lowest 20 % income | | | 1.7** | 1.2 - 2.5 | 1.7** | 1.2 - 2.6 |
| Lowest 20-40 % income | | | 1.6** | 1.1 - 2.2 | 1.6** | 1.1 - 2.2 |
| Middle 40-60 % income | | | 1.3+ | 1.0 - 1.8 | 1.3+ | 1.0 - 1.8 |
| Highest 60-80 % income | | | 1.2 | 0.9 - 1.6 | 1.2 | 0.8 - 1.6 |
| *Male sex* | 1.1+ | 1.0 - 1.3 | 1.1+ | 1.0 - 1.3 | 1.1+ | 1.0 - 1.3 |
| *Child received free school meals at age 5* | 1.3** | 1.1 - 1.6 | 1.3* | 1.0 - 1.5 | 1.3* | 1.0 - 1.6 |
| *Single parent status at birth* | 1.7*** | 1.4 - 2.1 | 1.6*** | 1.3 - 1.9 | 1.6*** | 1.3 - 1.9 |
| *Maternal age: under 16* | 2.6* | 1.1 - 6.4 | 2.4+ | 1.0 - 6.0 | 2.4+ | 1.0 - 6.0 |
| *Maternal age: 16 to 19* | 2.7*** | 2.0 - 3.5 | 2.3*** | 1.7 - 3.1 | 2.3*** | 1.7 - 3.1 |
| *Maternal age: 20 to 24* | 2.0*** | 1.6 - 2.5 | 1.8*** | 1.4 - 2.3 | 1.8*** | 1.4 - 2.3 |
| *Maternal age: 25 to 29* | 1.4*** | 1.2 - 1.7 | 1.4** | 1.1 - 1.7 | 1.4** | 1.1 - 1.7 |
| *Maternal age: 35 to 39* | 1.0 | 0.8 - 1.3 | 1.1 | 0.8 - 1.4 | 1.1 | 0.8 - 1.4 |
| *Maternal age: under 16* | 1.0 | 0.6 - 1.7 | 1.1 | 0.7 - 1.8 | 1.1 | 0.6 - 1.8 |
| *Number of siblings: 1* | 1.4*** | 1.2 - 1.7 | 1.3** | 1.1 - 1.6 | 1.3** | 1.1 - 1.6 |
| *Number of siblings 2* | 1.8*** | 1.4 - 2.2 | 1.5*** | 1.2 - 2.0 | 1.6*** | 1.2 - 2.0 |
| *Number of siblings 3* | 2.3*** | 1.7 - 3.1 | 2.0*** | 1.4 - 2.7 | 2.0*** | 1.4 - 2.7 |
| *Number of siblings 4 or more* | 3.2*** | 2.1 - 4.8 | 2.6*** | 1.7 - 3.9 | 2.6*** | 1.7 - 3.9 |
| | | | **Obesity** | | | |
| Most deprived 20 % IMD | 1.8*** | 1.5 - 2.1 | | | 1.6*** | 1.3 - 1.9 |
| Most deprived 20-40 % IMD | 1.7*** | 1.4 - 2.0 | | | 1.5*** | 1.3 - 1.8 |
| Middle 40-60 % IMD | 1.4*** | 1.1 - 1.7 | | | 1.2* | 1.0 - 1.5 |
| Least deprived 60-80 % IMD | 1.3** | 1.1 - 1.6 | | | 1.3* | 1.0 - 1.5 |
| Lowest 20 % income | | | 1.6*** | 1.3 - 2.0 | 1.4** | 1.1 - 1.7 |
| Lowest 20-40 % income | | | 1.7*** | 1.4 - 2.0 | 1.5*** | 1.2 - 1.8 |
| Middle 40-60 % income | | | 1.4*** | 1.2 - 1.7 | 1.3** | 1.1 - 1.5 |
| Highest 60-80 % income | | | 1.3** | 1.1 - 1.5 | 1.2* | 1.0 - 1.5 |
| *Male sex* | 0.9 | 0.9 - 1.0 | 0.9 | 0.9 - 1.0 | 0.9 | 0.9 - 1.0 |
| *Child received free school meals at age 5* | 1.1+ | 1.0 - 1.3 | 1.2+ | 1.0 - 1.3 | 1.1 | 1.0 - 1.3 |
| *Single parent status at birth* | 1.0 | 0.9 - 1.2 | 1.0 | 0.9 - 1.1 | 1.0 | 0.9 - 1.1 |
| *Maternal age: under 16* | 0.6 | 0.2 - 2.0 | 0.5 | 0.1 - 2.0 | 0.5 | 0.1 - 1.9 |

| | | | | | | |
|---|---|---|---|---|---|---|
| *Maternal age: 16 to 19* | 1.0 | 0.8 - 1.2 | 0.9 | 0.7 - 1.2 | 0.9 | 0.7 - 1.1 |
| *Maternal age: 20 to 24* | 1.1 | 1.0 - 1.3 | 1.1 | 0.9 - 1.2 | 1.0 | 0.9 - 1.2 |
| *Maternal age: 25 to 29* | 1.1 | 1.0 - 1.2 | 1.1 | 0.9 - 1.2 | 1.0 | 0.9 - 1.2 |
| *Maternal age: 35 to 39* | 1.1 | 0.9 - 1.2 | 1.1 | 0.9 - 1.3 | 1.1 | 0.9 - 1.3 |
| *Maternal age: under 16* | 1.1 | 0.8 - 1.5 | 1.2 | 0.9 - 1.6 | 1.2 | 0.9 - 1.6 |
| *Number of siblings: 1* | 1.0 | 0.9 - 1.1 | 0.9 | 0.8 - 1.0 | 0.9 | 0.8 - 1.0 |
| *Number of siblings 2* | 1.0 | 0.9 - 1.2 | 0.9 | 0.8 - 1.1 | 0.9 | 0.8 - 1.1 |
| *Number of siblings 3* | 1.1 | 0.9 - 1.4 | 1.0 | 0.8 - 1.3 | 1.0 | 0.8 - 1.3 |
| *Number of siblings 4 or more* | 1.1 | 0.8 - 1.5 | 1.0 | 0.7 - 1.3 | 1.0 | 0.7 - 1.3 |
| Observations | 15,367 | | 15,367 | | 15,367 | |

*Note: Maternal age at 9 months (ref: 30 to 35); Number of siblings (ref: none) *** p<0.001, ** p<0.01, * p<0.05, + p<0.10. Regression results using modified Poisson regression adjusted for the variables in italics.*

**Figure A4: Receiver operating characteristic curves based on the unadjusted regression model (table 1)**

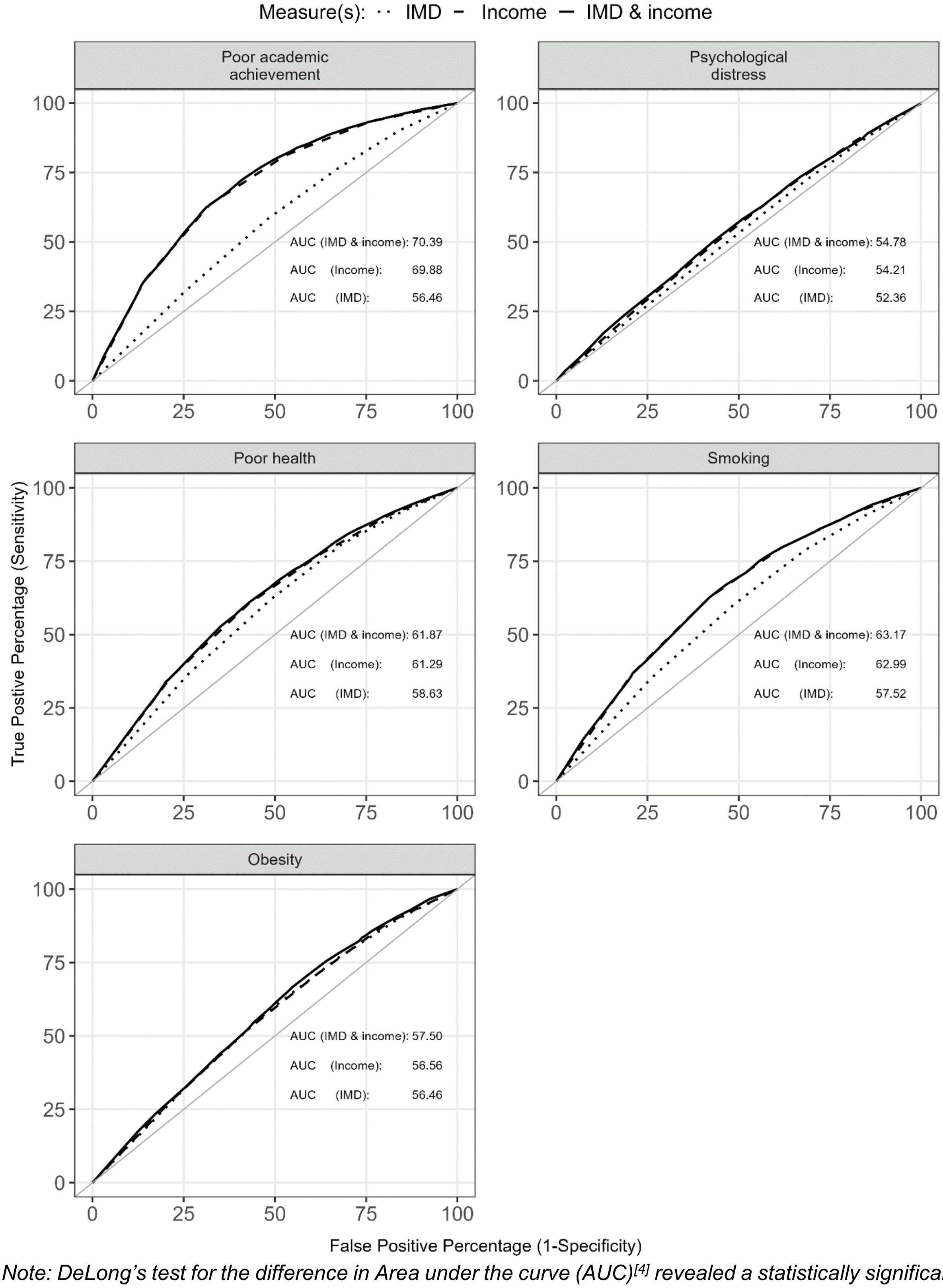


*Note: DeLong's test for the difference in Area under the curve (AUC)[4] revealed a statistically significant increase in AUC at 95% confidence level, when comparing a model with IMD quintiles as predictors (IMD) to a model with income quintiles as predictors (Income), for all of the adverse outcomes at age 17 except for obesity, for which the gain in AUC was not significant when using income vs. IMD quintiles as predictors. We found statistically significant increase in the AUC (95% confidence) when comparing a model with income quintiles as predictors (Income) and combined model (IMD & income) for all of the five adverse outcomes.*

**Figure A5: Receiver operating characteristic curves based on the adjusted regression model (table A4).**

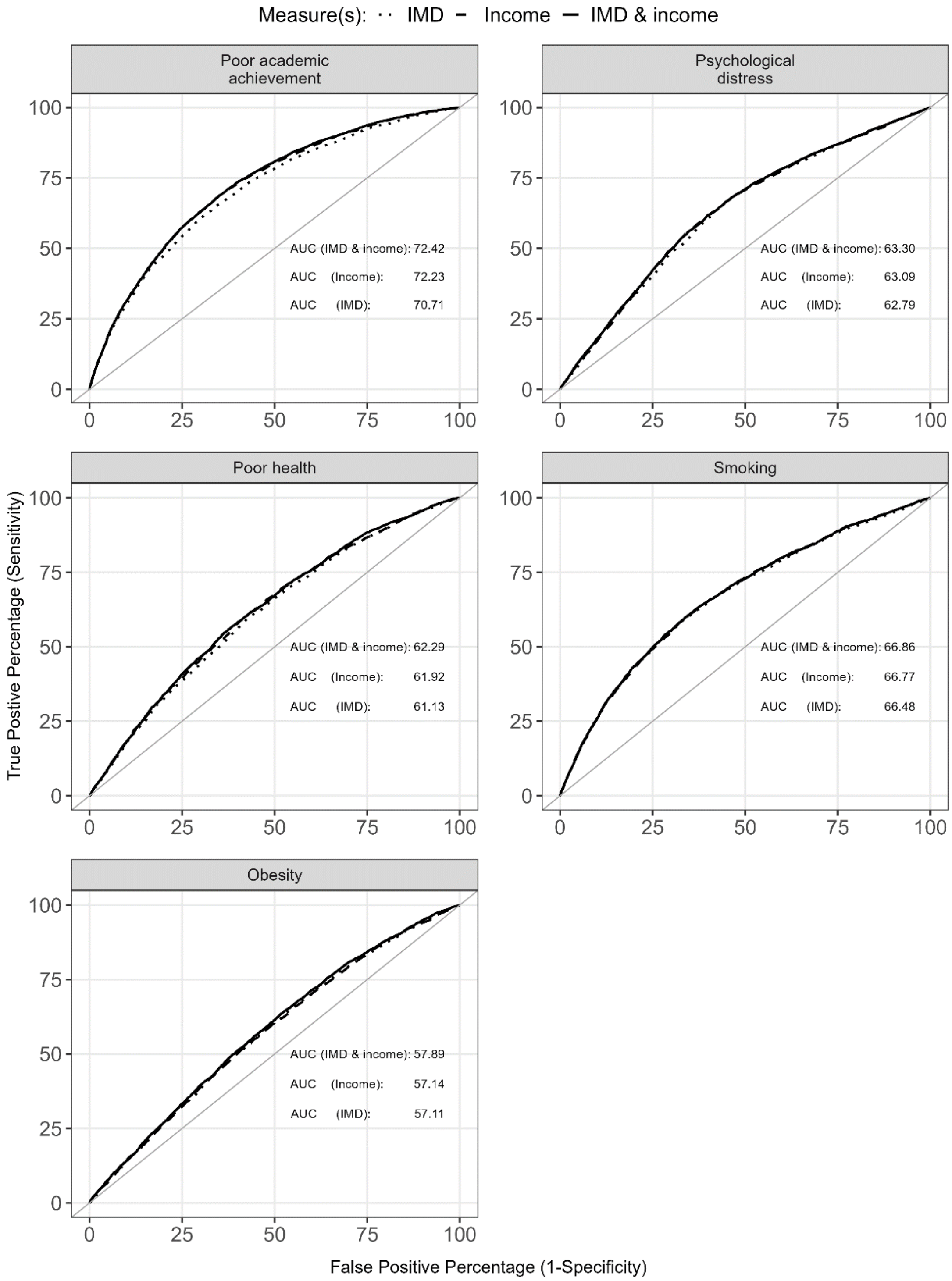


*Note: DeLong's test for the difference in Area under the curve (AUC)[4] revealed a statistically significant increase in AUC at 95% confidence level, when comparing a model with IMD quintiles as predictors (IMD) to a model with income quintiles as predictors (Income), for all of the adverse outcomes at age 17 except for obesity, for which the gain in AUC was not significant when using income vs. IMD quintiles as predictors. We found statistically significant increase in the AUC (95% confidence) when comparing a model with income quintiles as predictors (Income) and combined model (IMD & income) for all of the five adverse outcomes.*

4. DeLong, E.R., DeLong, D.M. and Clarke-Pearson, D.L., 1988. Comparing the areas under two or more correlated receiver operating characteristic curves: a nonparametric approach. *Biometrics*, pp.837-845.
